# Vancomycin Exposure is Associated with Protection from Acute Graft-vs-Host Disease (aGVHD) Through Microbiome-dependent Mechanisms

**DOI:** 10.1101/2025.11.22.25339693

**Authors:** Safa Elzein, Jennifer Trannguyen, Racheal Wilkinson, Randa Elzein, Shins Jose, Peter Monaco, Angelico Mendy, Kavya Patel, Dani Tapp, Tara Mink, Kitty Tierney, Michael Preziosi, James Mason, Senu Apewokin

## Abstract

**Backgound:** *Enterococcu*s and *Akkermansia* species enhance immune function through T-cell dependent mechanisms. This suggests vancomycin exposure and consequently the relative abundance of *Enterococcus* may have repercussions on immune-related outcomes such as GVHD. To test this hypothesis we assessed the a) association between vancomycin exposure and aGVHD b) association between stool *Enterococcus* abundance and aGVHD c) in-vitro effect of vancomycin on T-cell function.

**Methods:** Two distinct cohorts were evaluated. 1) A “derivation” cohort comprising 46 patients where vancomycin exposure data was correlated with aGVHD 2) A “validation” cohort of 26 patients where metagenomic sequencing assessed the correlation between enterococcal abundance and aGVHD. Additionally, ELISPOT assays evaluated the impact of vancomycin exposure on T-cell function.

**Results:** In the “derivation” cohort, aGVHD was associated with lower vancomycin exposure; (IQR 4.5 days) compared to (IQR 10.5 days) in patients without aGHVD (p-value 0.0072). Among five out of fourteen patients who developed aGVHD in “validation” cohort, significant genus enrichment for *Enteroccocus* and *Akkermansia* (5.75 and 7.59 log2fold change resp. p<0.05) was noted. Elispot assays demonstrated that T-cell exposure to vancomycin did not affect function.

**Conclusions:** Vancomycin exposure is associated with protection from aGVHD through microbiome-intrinsic processes but not direct impairment of T-cell function.

## INTRODUCTION

Hematopoietic stem cell transplantation (HSCT) stands as a well-established treatment modality for hematological malignancies and immune diseases. However, the development of Graft vs Host Disease (GVHD) undermines its success, and remains a significant contributor to post-HSCT mortality. The prevalent reliance on immunosuppression for GVHD treatment poses a noteworthy risk, amplifying susceptibility to infectious complications and infection-related mortality. In order to forge ahead with innovative therapeutic and preventive approaches for GVHD, a comprehensive exploration of risk factors and their associated mechanisms becomes imperative. Several studies have highlighted a link between gut dysbiosis and GVHD development. ^1,2,3^ The use of broad-spectrum antibiotics in HSCT recipients has been reported to be protective against GVHD. Evidence to support this was derived from studies in which gut decontamination successfully reduced GVHD incidence. ^1,4,5,6^ Recent studies, however, have shown an association between broad spectrum antibiotic use and development of graft vs host disease (GVHD). ^7,8,9^ Albeit discordant, both associations signify an interplay between intestinal microbiota, immune homeostasis, and the development of GVHD. The descripancy in findings across studies could be attributable to the lack of consistency among the broad spectrum agents used; given that antibiotic effects can be class-specific and these classes have unique effects on the gut microbiome.^10^ Recents reports have attempted to identify these class-specific and organism specific effects. One such study concluded that antibiotics active against anaerobic bacteria are associated with increased risks of GVHD post-HSCT ^11^ Hidaka al. also reported that carbapenem use for greater than 7 days was a risk for intestinal GVHD (iGVHD) ^12^. While these studies have provided consistent associations between antibiotic use and GVHD, the mechanisms underlying these observations remain unclear. To help elucidate the potential underlying mechanism we conducted clinical studies and functional T-cell assays to evaluate he association between gut microbiota, vancomycin exposure, and aGVHD

## METHODS

### Study Design

A “derivation” cohort, where we conducted a case/control retrospective analysis of 46 adult patients (>18 years old) who underwent allogenic HCT at Scripps Green Hospital (SGH), San Diego, CA from April 2017 to May 2020 was established. For the “validation” cohort, we enrolled 26 patients where metagenomic sequencing was performed on baseline stool (prior to chemotherapy onset and use of prophylactic antibiotics) to assess the correlation between the relative abundance of relevant microbes and aGVHD development.

All study procedures were approved by the Scripps Green Hospital and University of Cincinnati (UC) IRB and informed consent was obtained from all participants. Patients with GVHD were considered cases and those without GVHD were controls. Demographics, cumulative antibiotic exposure, and exposure to each antibiotic class were assessed. Patients who had successful engraftments were included in the study. Patients who had graft failure or death before engraftment were excluded. Two patients did not engraft initially but were able to engraft after a second donor and were included in the study. GVHD was diagnosed clinically and/or pathologically. Antibiotics were grouped into carbapenem, quinolones and vancomycins. Other antibiotic classes were used too infrequently to perform meaningful statistical comparisons. For the studies antibiotic exposure was considered during peri-engraftment period (7 days prior to transplantation and up to 30 days after transplant date).

Antibiotic Prophylaxis protocol : Patients received Levofloxacin 500 mg daily starting day 0 until ANC > 500 for two consecutive days. Sulfamethoxazole and Trimethoprim (SMX/TMP or PJP alternative) is started day +21 in all regimens except for post-transplant cyclophosphamide as GVHD prophylaxis where it starts on day+30. Patients who received antibiotics active against *Enterococcus* during peri-engraftment were excluded in the “validation” cohort to avoid confounding,. Classic acute GVHD (aGVHD) was defined as cases presenting within 100 days of hematopoietic cell transplant (HCT) and displaying features of acute GVHD. Cumulative antibiotic exposure was defined as total days of antibiotic therapy; multiple antibiotics given on the same day were counted as multiple antibiotic days.

### Statistical Analysis

The primary goal was to assess the association between antibiotic exposure, specifically, duration of each antibiotic, cumulative antibiotic duration, and incidence of GVHD. The secondary end point was incidence of acute GVHD. The analyzed variables included age, gender, Hematopoietic cell transplantation (HCT)-specific comorbidity index (HCT-CI), primary malignancy, conditioning regimen, date of transplant, degree of HLA match, aGVHD and GVHD prophylaxis. Descriptive statistics were used to summarize data, including frequencies and percentages to summarize categorical data, means and standard deviations to summarize normally distributed continuous data, and medians and interquartile ranges (IQR) to summarize skewed continuous data. Proportions were compared using Chi-Square tests, and durations were compared using Mann Whitney U tests between individuals with or without GVHD. Associations between specific antibiotic durations and the overall number of antibiotic days were tested using linear regression. All analyses were conducted using R v. 3.5.3.

### Microbiome analysis

Stool was collected from patients using the Omnigene kit and stored at -80 degree celcius until processed. To process stools, DNA extraction was performed using 0.25g of crude stool samples with Power Fecal DNA Isolation Kit^®^ by VMO BIO^®^ per kit instructions. DNA concentration was measured using Qubit^®^. Amplified library generation was be performed with Nextgen adapters, and sequencing performed to obtain 150bp DNA paired end reads to a depth of 2.5G base pairs per sample using a Illumina NovaSeq sequencing machine. The following annotation pipeline was used for all normalized metagenome reads. Kneaddata was employed for trimming and sequence read quality control. Kraken2 was used in conjunction with a custom database that include all RefSeq bacterial, fungal, parasitic, and viral genomes as well as additional bacterial and fungal genome sequences from the National Center for Bioinformatics to determine quantitative genus and species. An exact sequence read match of k-mer length 31 was done by Kraken2 with reads assigned to the lowest common ancestor. Bracken was then used for abundance estimation. Processing and normalization of count data to the lowest number of total reads mapped among the samples was performed using, phyloseq, microbiome and microbiome explorer packages in R to give the relative abundance at both the genus and species level. For microbial diversity assessments estimated Inverse Simpson index was performed. To compare taxa between the groups of interest we used DESeq2. DESeq2 fits a negative binomial distribution and uses the Benjamini-Hochberg (BH) adjustment for multiple testing considerations.

### Functional T-cell assays

To assess T-cell function we performed an ELISpot assay which is an immunoassay used to quantify analyte-secreting cells.^13,14^ Interferon gamma secreted by T-cells are captured by specific antibodies immediately after secretion. T-cells isolated from a healthy donor using the a ROBOSEP instrument were cultured in a plate with membrane surfaces coated with an interferon gamma capture antibody and in the presence of varying concentrations of vancomycin. 100000 cells were plated per well and incubated for 48 hours followed by assessments for secreted interferon gamma using a detection antibody. The number of spots were then counted using ImmunoSpot software version 5.3. Comparative statistics were then performed using Graphpad Prism.

## RESULTS

Of a total of 193 HSCT patients (both allogenic and autologous) 2017 -2020 in Scripps Green Hospital. 133 underwent autologous HSCT. 43 underwent Allogenic HSCT. Out of the 46 allogenic HSCT patients, 28 developed aGVHD (cases) and 18 did not develop aGVHD (controls). Of these 52% were male, median HCT-CI was 2 and the mean age was 55.7. 64.58% had an HLA 10/10 match. Median HCT-CI of 2 vs 3 in patients with aGVHD and without aGVHD respectively. (Table 1,2). aGVHD was associated with exposure to longer total/cumulative days of antibiotics (median days of 39.00 vs 20.00, p= 0.0351). (Table 3). Vancomycin use was associated with decreased incidence of aGVHD despite longer duration of antibiotics in the vancomycin group. For each one day increase in vancomycin exposure, the total days on antibiotics on average increased by 3.255. There was higher frequency of cefepime use in vancomycin use group. In the “derivation” cohort, 28 out of the 46 patients developed aGVHD (60%). This is comparable to the national rate of 20-80%. ^15^ There was no significant difference in baseline characteristics of patients with and without aGVHD except for comorbidity index of 2 vs 3 in patients with aGVHD and without aGVHD respectively. aGVHD patients had longer exposure or cumulative days of antibiotics during periengraftment period (median days of 39.00 vs 20.00, p= 0.0351).

**Table 1.** Patient demographics for all patients in the ““derivation”” cohort.

| Demographics |  |  |
| --- | --- | --- |
|  | N | % |
| Age at Transplant: Mean, SD | 55.71 | 15.42 |
| HCT-CI: Median, IQR | 2 | 3 |
| Gender (Male) | 25 | 52.08% |
| Disease |  |  |
| AML | 23 | 47.92% |
| MDS | 10 | 20.83% |
| Other/Multiple | 15 | 31.25% |
| Prep: |  |  |
| Flu/Mel | 28 | 58.33% |
| BU/CY | 10 | 20.83% |
| Other/None | 10 | 20.83% |
| HLA 10/10 Match | 31 | 64.58% |
| Engrafted | 46 | 95.83% |
| GVHD Prophylaxis: tac/mtx | 19 | 39.58% |

**Table 2.** Patient Demographics by aGVHD diagnosis for all pateints in the “derivation” cohort.

| Variable | Patients with<br>aGVHD (N=28) | Patients without<br>aGVHD (N=18) | P-value |
| --- | --- | --- | --- |
| Age at Transplant | 58.79 (12.69%) | 50.1 (18.53%) | 0.095 |
| HCT-CI: Median, IQR | 2 (3%) | 3 (1.75%) | 0.0397 |
| Gender (male) | 13 (46.43%) | 12 (66.67%) | 0.297 |
| Disease: AML | 13 (46.43%) | 9 (50.00%) | >0.999 |
| Conditioning treatment:<br>Flu/Mel | 20 (71.43%) | 7(38.89%) | 0.060 |
| HLA 10/10 Match | 18 (64.29%) | 12 (66.67%) | >0.999 |
| GVHD prophylaxis: tac/mtx | 11 (39.29%) | 8 (44.44%) | 0.968 |

**Table 3.** Cummulative Days of antibiotic use by aGVHD diagnosis. Median days of antibiotic use for each deriviation cohort group (aGVHD and Non-aGVHD) and associated interquartile range (IQR) for antibiotic treatment within each category.

|  | aGVHD (N = 28) |  | No aGVHD (N = 18) |  |  |
| --- | --- | --- | --- | --- | --- |
|  | Median | IQR | Median | IQR | p-value |
| **Total Antibiotic Days** | **39.00** | **32.00** | **20.00** | **17.25** | **0.0351** |
| **Quinolones** | 13.00 | 9.75 | 10.00 | 7.75 | 0.8392 |
| **cefepime** | 0.00 | 6.25 | 0.00 | 7.00 | 0.6998 |
| **Vancomycin** | 0.00 | 4.50 | 6.50 | 10.50 | 0.0072 |
| **Bactrim** | 0.50 | 2.25 | 1.00 | 7.00 | 0.2077 |

Microbiome Analysis: To examine the impact of relevant bacterial genera on development of aGVHD we compared the intestinal microbial composition of baseline stools between patients who developed aGVHD and those who did not. To avoid the confounding effects of *Enterococcus* ablation following antimicrobial exposure we excluded all patients who received glycopeptides during egraftment period. Metagenomic analysis revealed that cases did not differ from controls in alpha and beta diversity since we noted noted no difference in alpha (Shannon and Simpson indices) and beta diversity between subjects that developed GVHD and those who did not Fig 1. We also noted a higher relative abundance of *Enterococcus* and *Akkermansia* genera in subjects who developed GVHD compared to those who did not (5.75 and 7.59 log2fold change respectively. p<0.05). These genera were of particularly interest given numerous reports of their association with immune response.

**Figure 1.**
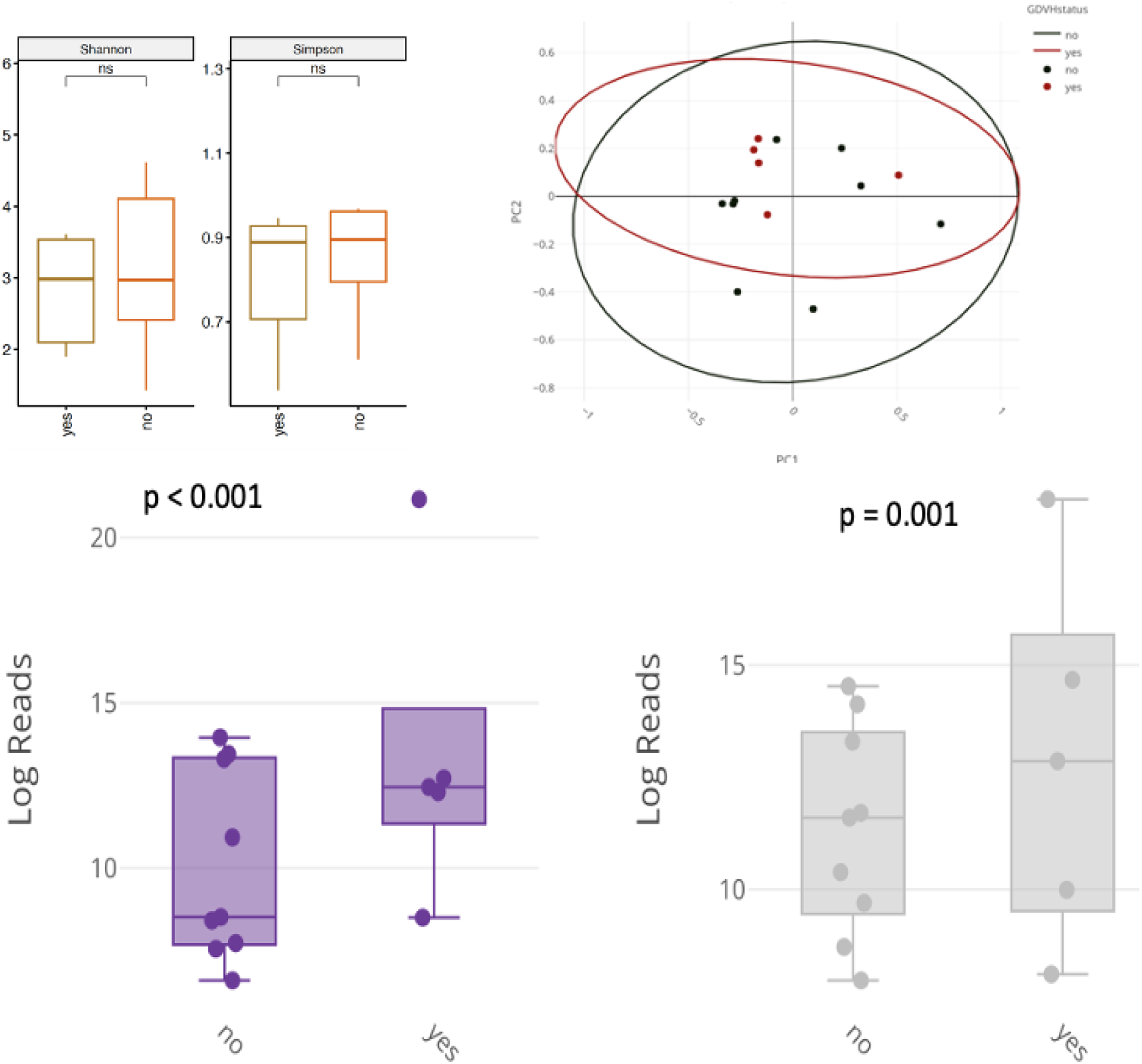
showing comparison of alpha and beta diversity measures between HSCT patients with aGVHD and without aGVHD in “derivation” cohorts (top panel) and comparison relative abundance of *Enterococcus* and *Akkermansia* (bottom panel)

Elispot assay: To establish that the effect of vancomycin exposure is through microbiome depent processes and not direct toxicity of vancomycin to T-cells, we performed elispot assays. We observed no difference in T-cell function, as depicted by the ability to secret interferon gamma, with varying doses of vancomycin (Fig. 2.).

**Figure 2.**
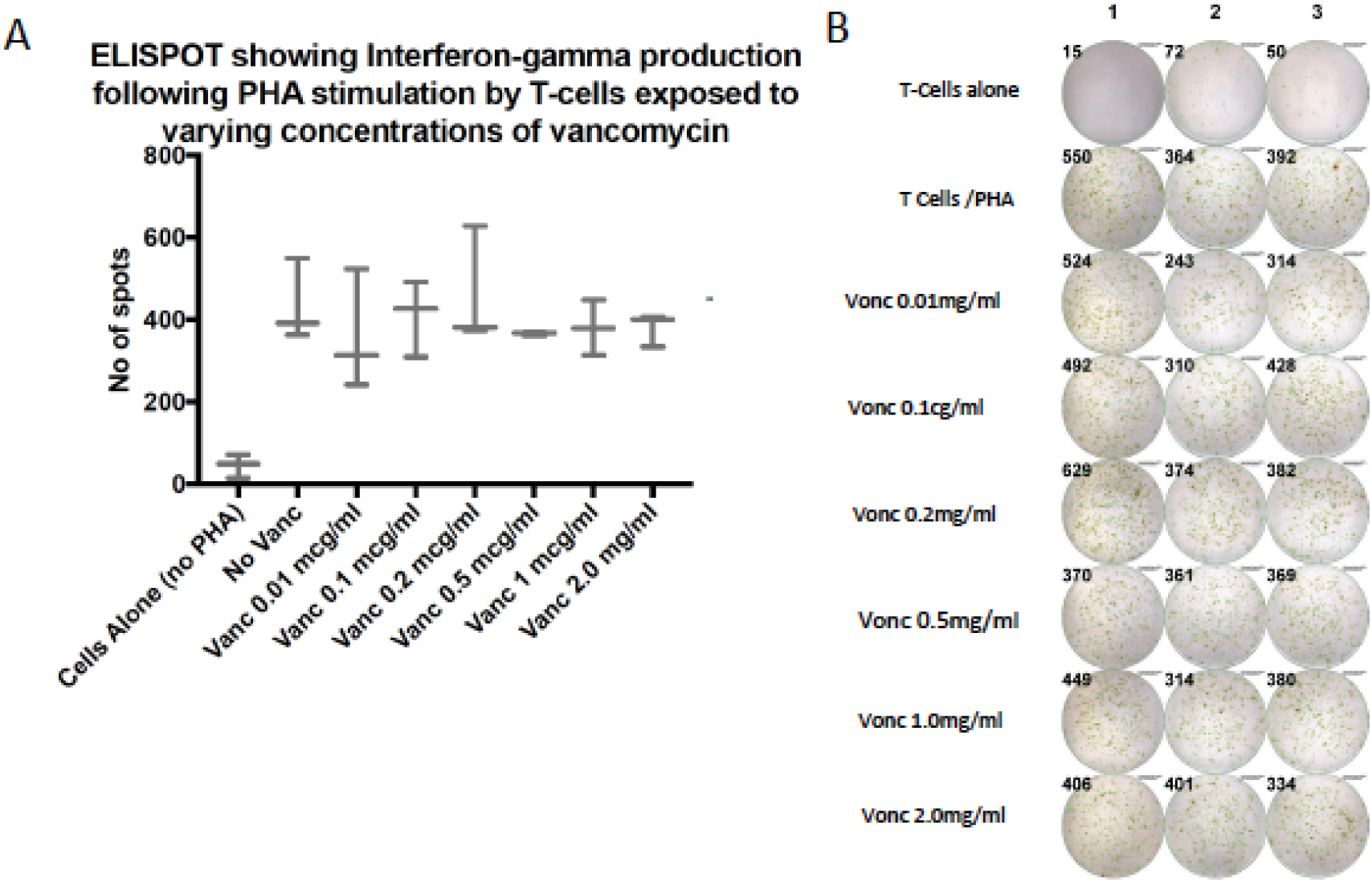
In vitro evaluation of T-cell function after exposure to varying concentrations of Vancomycin using ELISPOT assays

## DISCUSSION

Immune mediated mechanisms are major contributors to the toxicities observed after HSCT. To avoid this HLA matching remains a fundamental strategy that is employed in donor graft selection with hopes to minimize immune mediated toxicities.^16,17^ With increasing appreciation of influence of the gut microbiome on immune function there is a need to characterize the impact of the gut microbiome on immune mediated toxicities such as aGVHD. Consistent with our findings, Farowski et al. found that cumulative antibiotic exposure (>100 days) was associated with intestinal GVHD.^18^ Nishi et al demonstrated that exposure to antibiotics with activity against anaerobic bacteria, in particular, are associated with increased risks of aGVHD post-HSCT. In our study group, other antibiotic classes were used too infrequently to conduct meaningful statistical comparisons. A unique finding in our study was the protective effect of vancomycin. The use of Vancomycin was associated with decreased incidence of aGVHD despite longer duration of antibiotics in the vancomycin group. In landmark studies by Griffin et al. several *Akkermansia* and *Enterococcus* and a species were found to increase the efficacy of check point inhibitors by enhancing immune function through T cell dependent mechanisms.^19^ Building on this concept we hypothesized that the relative abundance of Enterococcus in the microbiome could impact and influence immune function and thereby impact immune related outcomes such as Graft vs Host Disease (GVHD). Consequently, drugs like vancomycin which preferentially eliminate *Enterococcu*s could have an impact on GVHD. A study by Smith et al indicated that microbiome changes were associated with CART cell efficacy. ^20^ They reported meropenem and vancomycin as some antibiotics that strongly affect T cell immunity and consequently related toxicties such as immune-cell associated neurologic syndrome. The authors also demonstrated several organisms including *Akkermansia* and *E. faecium* was associated with CART-cell efficacy. Our finding of a higher prevalence of *Akkermansia* in aGVHD is consistent with these finding and represents an intriguing observation given recent associations with enhanced immunity. It is a strict anaerobe utilizes mucin as its sole source of carbon and nitrogen elements. ^21^ Mucin forms a physical and chemical barrier that protects the host from pathogenic luminal bacteria.^22,23,24^ Thus the higher prevelance in patients aGVHD raises the possibility that it mediates its effects in immunity through degradation of the mucin layer and providing improved accessibility of immune cells in the lamina propia to luminal microrganisms Mucin degradants are also reported to regulate host immune system through signals such as tumour necrosis factor alpha (TNF-a), interferon gamma (INF-c), interleukin-10 (IL-10) and IL-4. ^25,26^

*Enterococcus* has been reported to be associated with the development of GVHD and has been tied to lactose availability..^27,28^ Our observations that *Enterococcus* was more abundant in the microbiome profiles of aGVHD patients is consistent with these findings. Since vancomycin has potent anti-enterococcal activity, our findings of protection from aGVHD following vancomycin exposure implicates reduction in the relative abundance of *Enterococcus* as a possible explanation. Conversely vancomycin has also been shown to have direct toxicity to many cell types thus to ascertain whether vancomycin has a direct impact on T-cell, we assessed for a vanc dose response effect on IFN gamma production by T cell. ^29,30^ Our results indicate that vancomycin did not cause direct impairment of T-cell function and alludes to microbiome mediated effects

In conclusion, Our study indicates that exposure to vancomycin is associated with a protective effect against acute graft-versus-host disease (aGVHD) through microbiome-intrinsic processes, rather than direct impairment of T-cell function. These results contribute to a deeper understanding of the intricate interplay between antibiotic exposure, gut microbiota, and immune outcomes, offering insights into potential therapeutic strategies for aGVHD prevention

## Data Availability Statement

All sequence data will be made publicly available

## Author Contributions Statement

**Safa Elzein**, (SE) Jennifer Trannguyen (JT), Racheal Wilkinson (RW), Randa Elzein (RE), Tana Mink (TM), Shins Jose (SJ), Peter Monaco (PM), Angelico Mendy (AM), Kavya Patel (KP), Dani Tapp (DT), Kitty Tierney (KT), Michael Preziosi (MP), James Mason (JM) and Senu Apewokin (SA)

SE,SA. Conceived /conceptualized study and planned the experiments. JT and SA, carried out the experiments. JT, RW, and PM. contributed to sample preparation. SA1, JR and RW collected clinical data, SE, JT, RW, RE., TM, SJ, AM, KP, DT, MC. KT MP, JM,SE and SA contributed to the interpretation of the results. SE and SA2 took the lead in writing the manuscript. SA peformed microbiome and statistical analysis. All authors provided critical feedback and helped shape the research, analysis and manuscriptAll authors have reviewed the manuscript. contributed to the final manuscript

## Additional Information

*SA and AM have work funded by The National Institutes of Health

